# Repeatability and agreement of central corneal thickness measurements with a new handheld non-contact pachymeter

**DOI:** 10.1101/2023.04.19.23288788

**Authors:** John G Lawrenson, Simranjit Gill, Isra Masuid, Fardip Rashid

**Affiliations:** Centre for Applied Vision Research, School of Health and Psychological Sciences, City University of London, London UK; Moorfields Eye Hospital NHS Foundation Trust, London UK; Royal Berkshire NHS Foundation Trust, Reading UK

**Author notes:** **Author contributions: John G Lawrenson:** Conceptualization (equal); data curation (equal); formal analysis (equal); investigation (equal); methodology (equal); project administration (equal); writing – original draft (equal). **Simranjit Gill:** Conceptualization (equal); investigation (equal); methodology (equal); writing – review and editing (equal). **Isra Masuid:** Conceptualization (equal); investigation (equal); methodology (equal); writing – review and editing (equal). **Fardip Rashid:** Conceptualization (equal); investigation (equal); methodology (equal); writing – review and editing (equal). **Ethics approval statement:** The study conformed to the tenets of the Declaration of Helsinki and was approved by the School of Health and Psychological Sciences Research and Ethics Committee (REC reference: ETH2021-1765). **Patient consent statement:** Written informed consent was obtained from all participants.

**Keywords:** central corneal thickness, pachymetry, agreement, precision

## Abstract

**Purpose:** To compare the repeatability of central corneal thickness (CCT) measurements taken with a new hand-held pachymeter (Occuity PM1 Pachymeter) and to assess its agreement with ultrasound biometry and two commercially available optical biometers in participants with normal eyes.

**Methods:** Three consecutive CCT measurements of the right eye of 105 participants with normal corneas were acquired by the PM1 pachymeter, Lenstar LS900 and Oculus Pentacam HR in a random order. This was followed by three measurements with a hand-held ultrasound pachymeter (UP) (Pachmate-2). Repeatability and the repeatability limit were calculated with each device and Bland-Altman limits of agreement (LoA) were determined for the PM1 pachymeter compared to the other devices.

**Results:** The mean CCT (±SD) was 551.04±33.43 μm, 558.62±31.46 μm, 549.41±31.00 μm, and 539.73±29.50 μm for the PM1 pachymeter, UP, Lenstar and Pentacam, respectively. The repeatability limits (expressed as the within subject SD for repeat measurements) were 14.02, 13.68, 4.99 and 9.90 μm respectively. The closest agreement was between the PM1 and Lenstar (mean difference = 1.63 μm with LoA 10.72 μm below and 13.97 μm above the readings obtained with the Lenstar. The PM1 underestimated CCT compared to UP (mean difference = 7.58 μm, LoA 24.63 μm below and 9.47 μm above UP. The agreement was lowest between the PM1 and Pentacam (mean difference= -11.30 μm, LoA between 4.29 μm and 26.89 μm).

**Conclusions:** The PM1 pachymeter shows excellent precision for CCT measurements across a range of corneal thicknesses in normal eyes and provides a safe and easy to use alternative to ultrasound pachymetry.

**Key points:**

- Measurement of central corneal thickness (CCT) is an important clinical measurement in the diagnosis and management of glaucoma and certain corneal dystrophies, it also provides useful information prior to refractive surgery
- The Occuity PM1 pachymeter is a new hand-held non-contact perimeter that uses confocal technology to measure CCT. The device showed excellent precision for CCT measurements across a range of corneal thicknesses in normal eyes
- The PM1 pachymeter provides a safe and easy to use alternative to ultrasound pachymetry that could facilitate an increased uptake of pachymetry in routine optometric practice

## Introduction

Assessment of central corneal thickness (CCT) is a key measurement for many aspects of clinical practice. It is widely accepted that intraocular pressure (IOP) readings are overestimated in eyes with a thicker cornea and underestimated with thinner cornea. [1] CCT also provides valuable information in risk profiling patients with ocular hypertension (OHT) and suspect glaucoma, [2] since a thinner CCT is associated with a greater risk of conversion of OHT to glaucoma and increased risk of glaucoma progression. [3] Evaluation of CCT is also useful in the diagnosis and monitoring of corneal dystrophies [4] and prior to refractive surgery, to ensure that the thickness of the residual corneal bed after surgery is sufficient to minimise the risk of postoperative corneal ectasia. [5] Despite its clinical importance, there is evidence that corneal pachymeters are not widely available to community optometrists [6,7] and that many optometrists lack confidence in performing and interpreting pachymetry results.[8]

A-mode ultrasound (A-scan) biometry (UB) has historically been the reference standard for measurement of CCT in routine ophthalmic practice. The method relies on the transit time of the ultrasound pulse from the transducer to the posterior corneal surface and back again. [1] Modern hand-held ultrasound pachymeters are light, portable, quick to use and relatively inexpensive. As a result, these contact devices are the instrument of choice in busy ophthalmology clinics.

A number of non-contact optical biometers are commercially available that integrate corneal topography with repeatable and accurate measurements of CCT and a range of ocular biometry measurements for intraocular lens power calculations and to aid myopia management. [9] However, these sophisticated instruments are expensive and lack portability.

The Occuity PM1 pachymeter is a new hand-held non-contact pachymeter that uses confocal scanning technology. The principle of measurement is that a tightly focussed beam of laser light is directed into the eye and the focal point scanned across the cornea. Whenever the focal point meets a surface, the anterior and posterior corneal surfaces in this case, a bright reflection is seen on the confocal receiver. By measuring the separation between the reflections within the scan, the corneal thickness is determined, via a correction factor to account for the refractive index of the cornea.

The aim of this study was to compare the repeatability of measurements taken with the PM1 Pachymeter and to assess the level of agreement with ultrasound biometry and two commercially available optical biometers in participants with normal eyes.

## Methods

One hundred and five eyes of 105 adult subjects aged 18 years and over, were prospectively enrolled into the study. Exclusion criteria included existing or previous ocular surface pathology, history of ocular surgery (including refractive surgery) and inability to fixate on a target e.g. in strabismus. Subjects with refractive error were not excluded; however, contact lens wearers were instructed not to wear their lenses on the day of testing. The study received ethical approval from the School of Health and Psychological Sciences Research and Ethics Committee (REC reference: ETH2021-1765) and was conducted in accordance with the Declaration of Helsinki. In compliance with UK medical device regulations, the clinical investigation also received a letter of ‘no objection’ from the MHRA (reference: CI/2022/0063/GB). All participants provided written, informed consent before taking part. A payment of £30 was provided as compensation for participation.

Three consecutive CCT measurements were taken from the right eye of each subject, using three optical (non-contact) biometers (PM1 pachymeter (Occuity Ltd); Lenstar LS 900 (Haag-Streit Ltd) and Pentacam (OCULUS Optikerate GmbH). The order of testing with these instruments was randomised. This was followed by three measurements taken with a hand-held ultrasound pachymeter. Ultrasound pachymetry was performed last due to the potential effect of topical anaesthesia on CCT.

### PM1 pachymeter

To take a measurement with the PM1 pachymeter (figure 1), the operator touches the screen to put the PM1 into scanning mode and holds the device up to the patient’s eye. The PM1 utilises a novel video alignment system to ensure that the measurement is taken at the corneal centre. Once aligned the PM1 automatically begins measuring, capturing approximately 200 scans per second, and then displays the average of these measurements.

**Figure 1.**
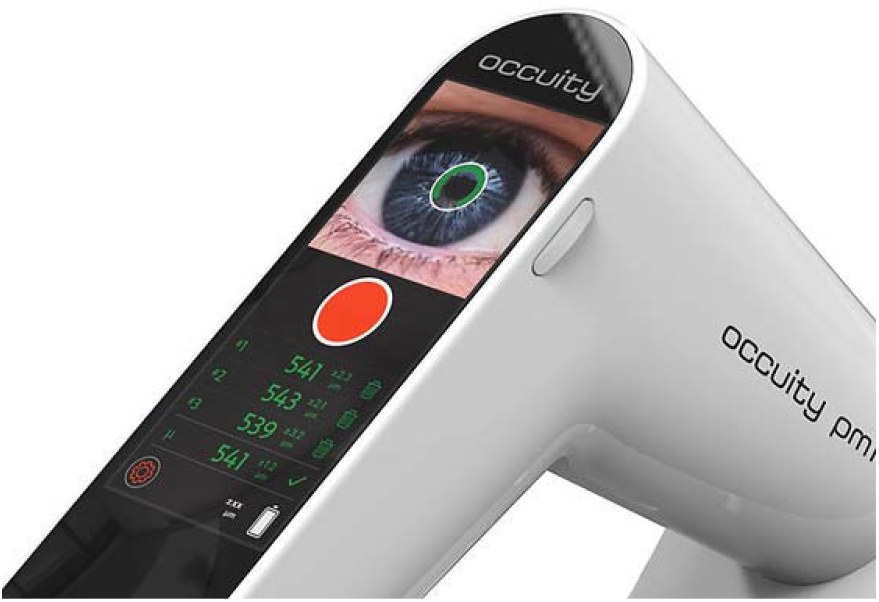
Display of the PM1 pachymeter

### Lenstar LS900

The Lenstar uses the principle of optical low coherence interferometry and uses an 820 nm superluminescent diode to measure several eye thickness and length measurements. Participants were seated with their heads stabilised using a chin rest and brow bar and were asked to fixate on the internal red fixation light while the measurements were taken. The instrument was aligned using the image of the eye on the computer monitor. Five measurements were taken as recommended by the manufacturer to calculate the mean CCT.

### Oculus Pentacam HR

The Pentacam utilises a rotating Scheimpflug camera, which captures images of a 475 nm monochromatic slit of light that illuminates the cornea. The 25-picture scan mode was used, consisting of 25 pictures per second. Using data from these images, the system calculates a 3D model of the anterior segment. Images were captured with the subjects seated and their head stabilised with a chin rest and brow bar. CCT was extracted from the ‘Pachymetry Map’, which reports thickness values at the corneal apex, the pupil centre and thinnest point. The CCT at the pupil centre was used in the current analysis.

### Ultrasound biometry

Consecutive CCT measurements were then taken with an ultrasound pachymeter (Pachmate 2 (DGH Technology Inc). A drop of the topical anaesthetic, 0.5% proxymetacaine hydrochloride (Chauvin Pharmaceuticals Ltd), was instilled into the conjunctival sac to anaesthetise the cornea and the probe was positioned on the central cornea perpendicular to the corneal surface. With the device in ‘Continuous Averaging Mode’, the pachymeter makes 25 measurements and provides the mean CCT value.

Measurements with all biometers were taken within a short time of each other during the period 10.00 to 17.00 hours by experienced examiners, who were masked to the other test results. Total test time was 30-40 minutes.

### Statistical analysis

Statistical analyses were performed with MedCalc for Windows, version 19.4 (MedCalc Software, Ostend, Belgium) using methods that conformed to established guidance on conducting agreement and precision studies. It is recommended that sample sizes for agreement studies should be at least 100 subjects, based on the accuracy of the estimates of the limits of agreement [10,11]. Descriptive statistics were calculated as the mean CCT and standard deviation (SD) for each device.

Repeatability was defined as the variability in repeated measurements with each instrument when other factors are assumed to be constant (examiner, calibration and time between measurements). To evaluate repeatability, a repeat measures analysis of variance (ANOVA) was performed to determine the within subject standard deviation (*S*_*r*_). The 95% confidence interval (CI) around *Sr* represents the repeatability limit (*r*), within which 95% of the measurement lie (calculated as 1.96 √2 x *Sr*).

To assess agreement between devices, the Bland Altman limits of agreement method, which plots the differences in the measurement between two devices against their mean, was used. Agreement is summarised by the bias, i.e., the mean difference and standard deviation of the differences. The limits of agreement (LoA) are defined as the mean difference plus and minus 1.96 times the standard deviation of the differences.

## RESULTS

The right eye of 105 subjects, 49 (46.7%) female, with mean age 41.3 ±14.5 years (range 19–74 years) and ethnicity 82.9% white, 9.5% Asian, 2.9% black, 4.8% other was measured with each biometer.

The mean, SD, repeatability (Sr) and repeatability limit (r) of the PM1, Lenstar, Pentacam and ultrasound pachymeter for the measurement of CCT are shown in table 1. The repeatability of the PM1 was approximately five microns and similar to the ultrasound pachymeter with comparable repeatability limits of approximately 14 μm (the likely limits within which 95% of measurements should occur). Repeatability with the Lenstar and Pentacam were better than both hand-held devices (approximately 2 μm). The Lenstar showed the narrowest repeatability limit (approximately 5 μm).

**Table 1.** The mean, SD, repeatability (Sr) and repeatability limit of CCT for each biometer

| Device | Mean CCT ±SD $\mu\text{m}$ | Repeatability (Sr) | Repeatability limit (r) |
| --- | --- | --- | --- |
| PM1 | 551.04±33.43 | 5.06 | 14.02 |
| UP | 558.62±31.46 | 4.94 | 13.68 |
| Lenstar | 549.41±31.00 | 1.80 | 4.99 |
| Pentacam | 539.73±29.50 | 2.13 | 9.90 |

*Abbreviations: CCT= central corneal thickness; UP=ultrasound pachymetry*

In terms of agreement, table 2 shows the mean difference, SD and 95% LoA (with 95% CIs) for comparisons between the PM1, ultrasound pachymeter, Lenstar and Pentacam. All mean differences were statistically significant. The closest agreement was between the PM1 and Lenstar with a mean difference of -1.63 μm and relatively narrow LoA’s with the PM1 giving readings up to 10.72 μm below and 13.97 μm above the readings obtained with the Lenstar. Overall, the PM1 underestimated the CCT compared to ultrasound pachymetry, mean difference 7.58 μm, with LoA up to 24.63 μm below and 9.47 μm above the ultrasound pachymetry. The agreement was lowest between the PM1 and Pentacam (mean difference=-11.30 μm, LoA between 4.29 μm and 26.89 μm). Figures 2-4 show the Bland-Altman plots for the three pairwise comparisons.

**Table 2.** The mean difference, SD, p value, limits of agreement (LoA) with 95% CIs between the PM1 and the other three biometers.

| Comparison | Mean difference in CCT (95% CI) | p value | Lower 95% LOA (95% CI) | Upper 95% LOA (95% CI) |
| --- | --- | --- | --- | --- |
| UP vs PM1 | 7.58 (5.90 to 9.27) | <0.0001 | -9.47 (-12.35 to -6.58) | 24.63 (21.75 to 27.52) |
| Lenstar vs PM1 | -1.63 (-2.85 to -0.41) | 0.0093 | -13.97 (-16.06 to -11.88) | 10.72 (8.63 to 12.80) |
| Pentacam vs PM1 | -11.30 (-12.84 to -9.76) | <0.001 | -26.89 (-29.53 to -24.25) | 4.29 (1.65 to 6.93) |

*Abbreviations: UP=ultrasound pachymetry*

**Figure 2.**
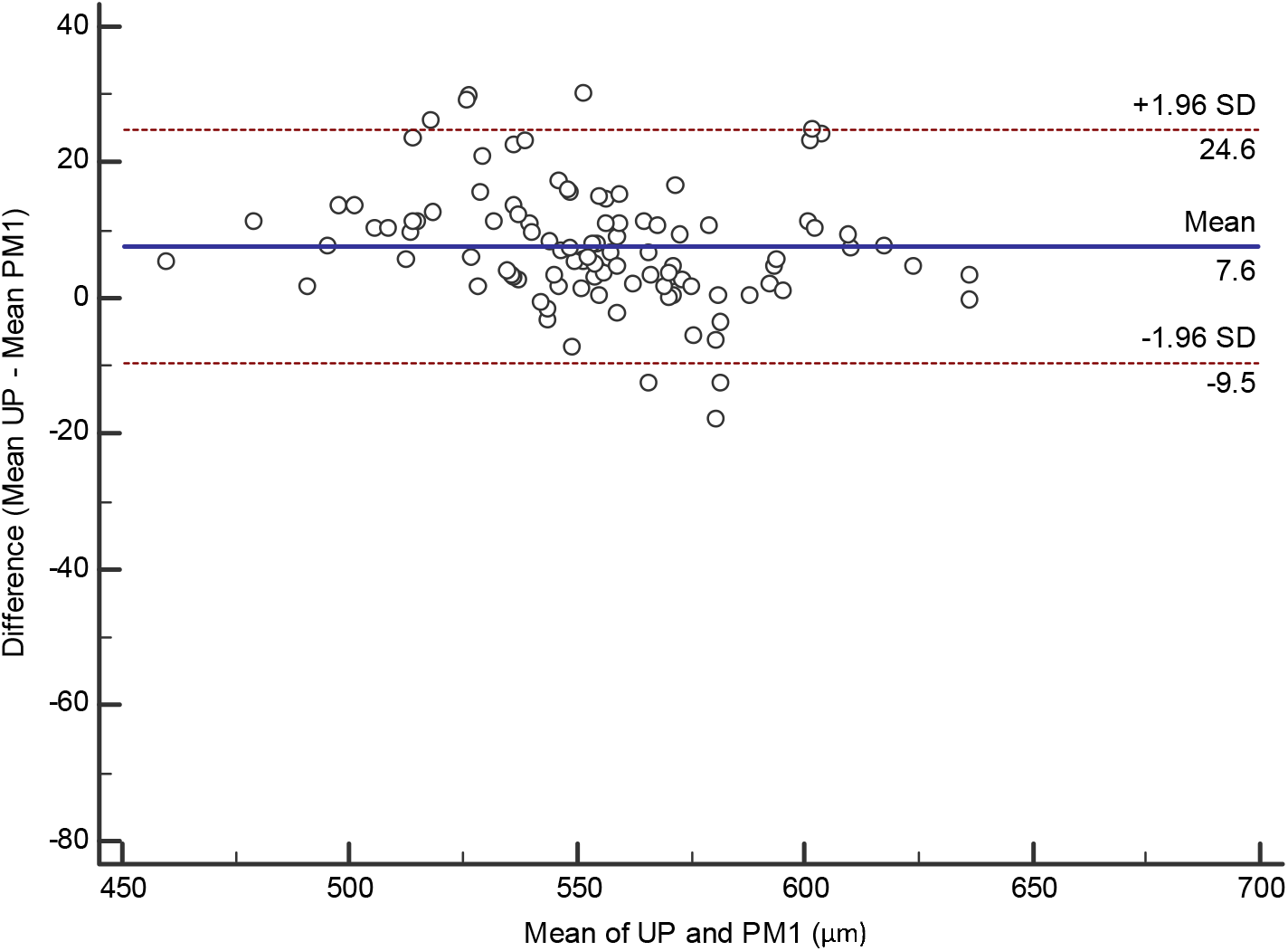
Bland-Altman plot showing the agreement between ultrasound biometry and the PM1 for CCT measurements. The solid line indicates the mean difference (bias), and the dotted lines indicate the 95% limits of agreement.

**Figure 3.**
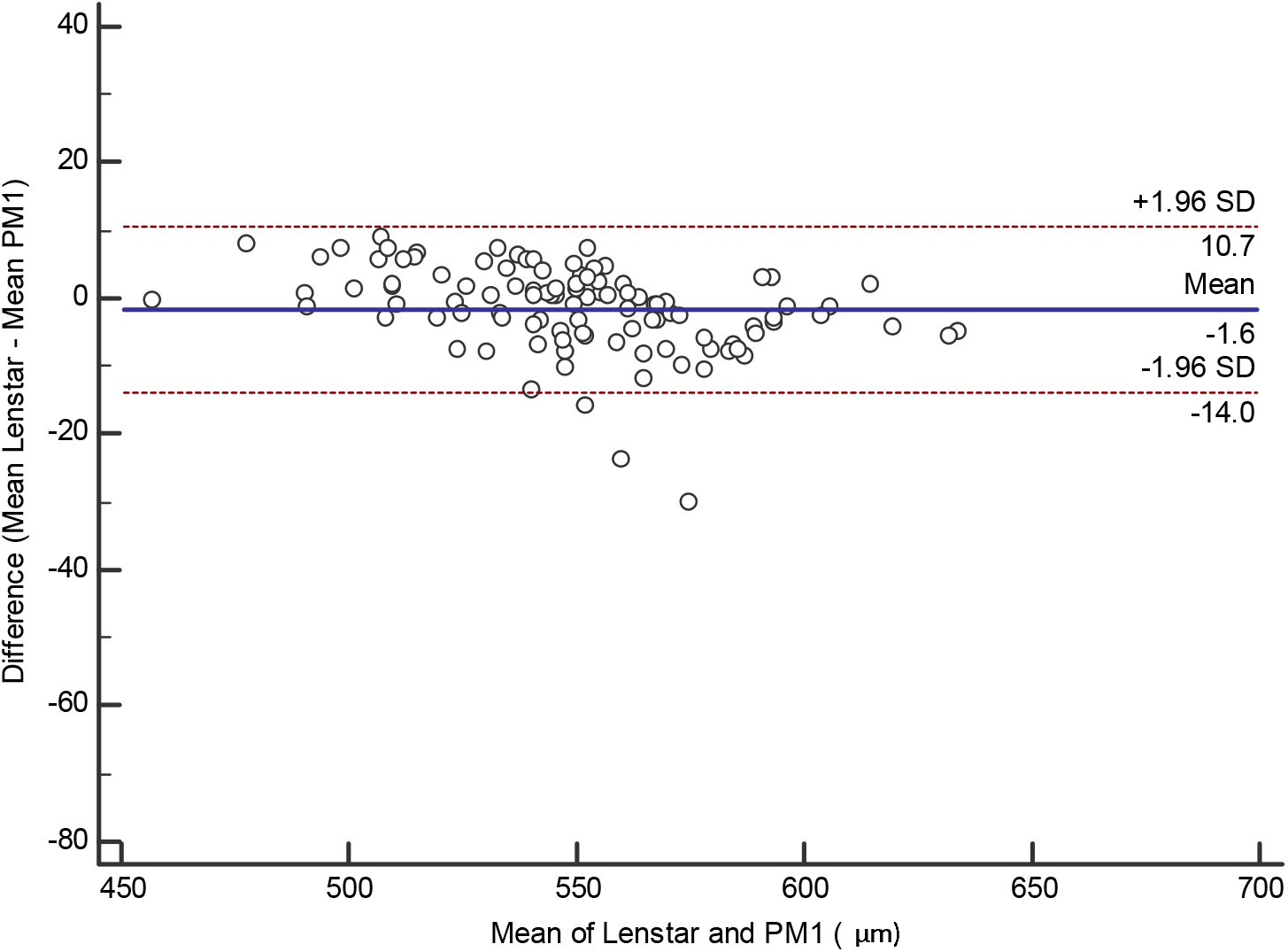
Bland-Altman plot showing the agreement between the Lenstar and the PM1 for CCT measurements. The solid line indicates the mean difference (bias), and the dotted lines indicate the 95% limits of agreement.

**Figure 4.**
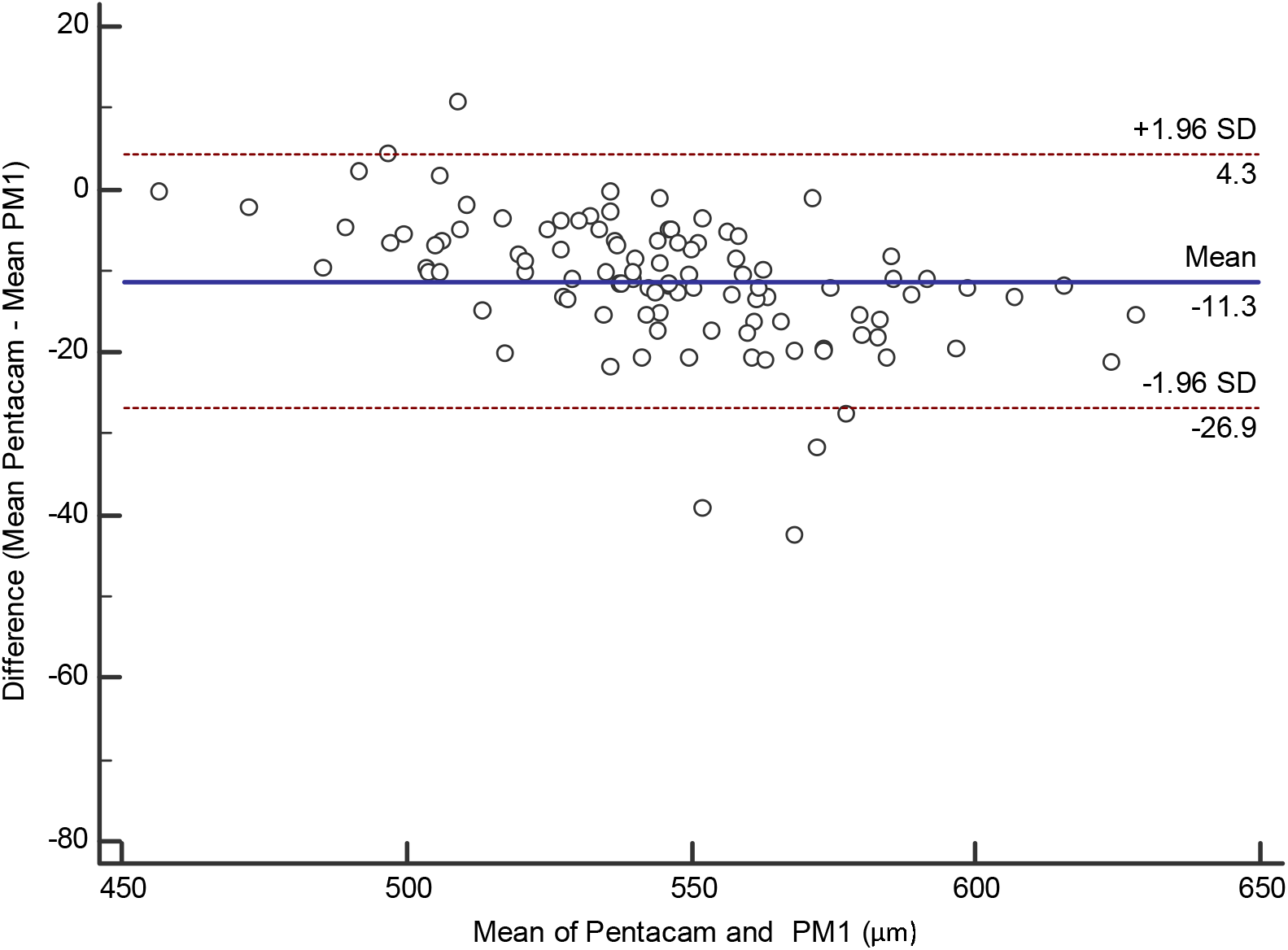
Bland-Altman plot showing the agreement between the Pentacam and the PM1 for CCT measurements. The solid line indicates the mean difference (bias), and the dotted lines indicate the 95% limits of agreement.

To illustrate the comparative agreement between devices, we plotted the difference between the overall mean CCT for all devices and the mean for each specific device against the overall mean (figure 5). The 95% limits of agreement were 4.3 μm above to 29.9 μm below the overall mean, with optical devices generally showing thinner CCT measurements compared with ultrasound pachymetry. Overall, the PM1 showed good agreement across the range of CCT measurements.

**Figure 5.**
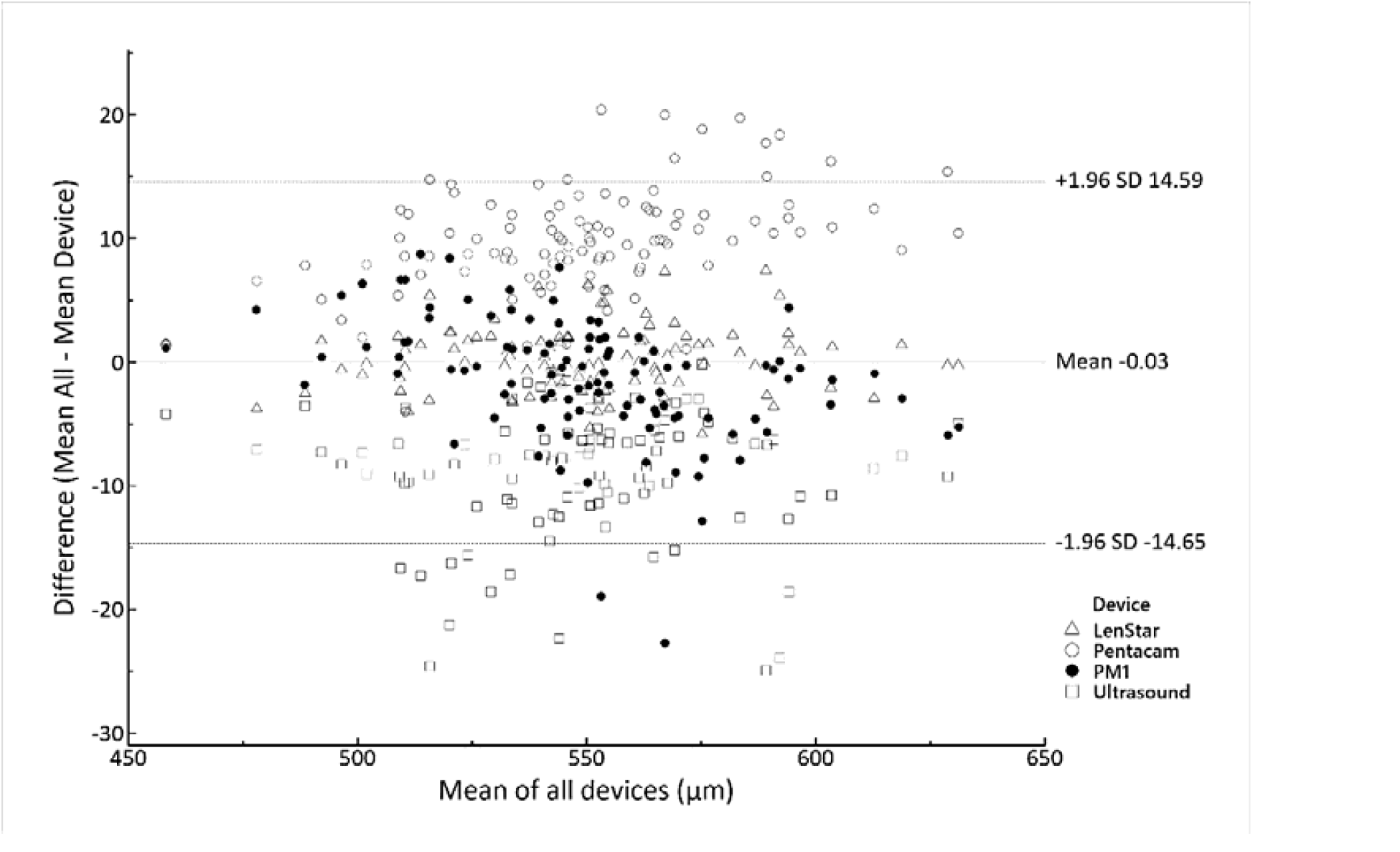
Bland-Altman plot showing the agreement between the mean measurement for each device against the mean CCT for all devices. The solid line indicates the mean difference (bias), and the dotted lines indicate the 95% limits of agreement. Measurements for the PM1 are highlighted.

## Discussion

Technological advances are transforming eye care. The influx of new instrumentation to measure various ocular biometric parameters has driven research into the validity of these measurements. [9] Questions that are important to the clinician include the accuracy of the measurements, which is usually defined by repeatability and reproducibility, [11] and how the measurements with the new instrument compare to the ‘gold standard’, to determine whether readings from different devices can be used interchangeably. [10]

The current study investigated the repeatability of a new optical pachymeter (Occuity PM1 Pachymeter) and compared its agreement with other commercially available devices in normal eyes. Ultrasound biometry is generally considered to be the reference standard for CCT measurements. Although measurements with ultrasound pachymeters have been shown to provide excellent inter and intra-observer agreement, [12-14] there are a number of drawbacks to this method. The contact of the biometer probe with the cornea requires topical anaesthesia, which causes discomfort and in some individuals may affect corneal thickness (due to corneal epithelial oedema) [15,16]. Furthermore, contact with the cornea increases the risk of cross-contamination and the accuracy of the measurement may depend on the experience of the user, as the probe needs to be aligned perpendicularly to the centre of the cornea.

We found a high level of repeatability for the PM1 pachymeter. The within subject standard deviation (repeatability) of three consecutive readings was approximately 5 μm, with a repeatability limit (range containing 95% of the measurements) of approximately 14 μm. This was similar to the repeatability of the ultrasound pachymeter. As expected, the Lenstar and Pentacam showed a higher degree of precision, with repeatability limits of approximately 5 and 10 μm respectively, confirming the results of earlier studies. [17-19]

The pairwise comparisons showed that measurements obtained with the PM1 pachymeter were slightly lower than ultrasound pachymetry, with a mean difference of 7.58 μm over a wide range of CCTs. Previous studies have similarly found that CCT measurements with ultrasound are slightly thicker than with optical biometers. [20-23] Although statistically significant, the small overall difference in CCT between the PM1 and ultrasound biometry is not clinically significant, however the 95% limits of agreement ranging from 9.47 μm above to 24.63 μm below would suggest that the two devices are not directly interchangeable and therefore the same device should be used when monitoring CCT over time. Similar results have also been found in agreement studies using optical coherence tomography (OCT), which is currently widely used for quantitative and qualitative evaluation of anterior segment structures in a variety of clinical settings. [24,25] Comparisons of OCT with ultrasound biometry have reported differences in mean CCT of up to 26 μm with limits of agreement of over 30 μm. [20,26,27]

There was closer agreement between the PM1 and Lenstar, with a mean difference of only 1.63 μm and narrower limits of agreement (13.97 μm above to 10.72 μm below the Lenstar). Measurements with these devices could be considered to be clinically interchangeable. By contrast, the PM1 generally showed higher CCT measurements (mean difference 11.3μm) than those taken from 25-picture scan with the Pentacam at the pupil centre. There was also greater variability (limits of agreement, 26.89 μm above to 4.29 μm below). Previous studies comparing the Pentacam with other optical biometers have reported similar levels of agreement. [18,19,27]

One of the main advantages of non-contact pachymeters is the reduced risk of cross-infection. Contact pachymetry has been identified as a possible infection risk as the majority of devices use non-disposable probes that require disinfection after use. Both the UK College of Optometrists and Royal College of Ophthalmologists have issued guidelines for infection control. [28,29] These recommend immersion of the pachymeter probe in freshly prepared sodium hypochlorite solution 1% (10,000 ppm of available chlorine) for at least 10 minutes, followed by rinsing in 3 changes of sterile water or saline for at least 10 minutes. A recent survey of pachymeter use and disinfection practice in UK ophthalmology units [30] identified significant variability in disinfection methods with only a few units following current professional guidance. The majority (66%) reported that they cleaned the pachymeter probe with an alcohol wipe between patients. Alcohol wipes are not 100% effective due to their inability to deactivate bacterial spores and 70% alcohol is ineffective against adenovirus and coronavirus. [31]

There are a few limitations to the current study. All participants had normal eyes and the performance of the PM1 pachymeter in eyes with ocular pathology or post LASIK was not assessed. Furthermore, all measurements were taken by the same group of experienced examiners and the results may not be generalisable to less experienced operators. This is particularly the case with ultrasound pachymetry, as all examiners were optometrists with a specialist qualification in glaucoma who were routinely performing this technique in their clinical practice.

In conclusion, the current study has demonstrated that the Occuity PM1 pachymeter shows excellent repeatability for CCT measurements in normal eyes. Although the measurements with the pachymeter closely agree with those obtained by ultrasound, as with the findings from earlier studies comparing other optical biometers to ultrasound, the measurements are not directly interchangeable, since the limits of agreement include clinically important differences. To the best of our knowledge, the PM1 is the first available hand-held non-contact pachymeter. With minimal training, the instrument is easy to use and the touch screen interface guides the operator to ensure correct alignment. As a result, the PM1 could be used by non-clinical staff and provides an opportunity for the measurement of CCT to become more firmly embedded in routine eyecare.

## Data Availability

All data produced in the present study are available upon reasonable request to the authors

## Acknowledgments

None

